# Supportive Care for Patient with Respiratory Diseases: An Umbrella Review

**DOI:** 10.1101/2020.04.13.20064360

**Authors:** Xufei Luo, Meng Lv, Xiaoqing Wang, Xin Long, Mengjuan Ren, Xianzhuo Zhang, Yunlan Liu, Weiguo Li, Qi Zhou, Yanfang Ma, Toshio Fukuoka, Hyeong Sik Ahn, Myeong Soo Lee, Zhengxiu Luo, Enmei Liu, Xiaohui Wang, Yaolong Chen, on behalf of COVID-19 evidence and recommendations working group

## Abstract

**Background:** Supportive treatment is an important and effective part of the management for patients with life-threatening diseases. This study aims to identify and evaluate the forms of supportive care for patients with respiratory diseases.

**Methods:** An umbrella review of supportive care for patient respiratory diseases was undertaken. We comprehensively searched the following databases: Medline, EMBASE, Web of Science, CNKI (China National Knowledge Infrastructure), Wanfang Data and CBM (SinoMed) from their inception to 31 March 2020, and other sources to identify systematic reviews and meta-analyses related to supportive treatments for patient with respiratory diseases including COVID-19, SARS, MERS and influenza. We assessed the methodological quality using the AMSTAR score and the quality of the evidence for the primary outcomes of each included systematic review and meta-analysis.

**Results:** We included 18 systematic reviews and meta-analyses in this study. Most studies focused on the respiratory and circulatory support. Ten studies were of high methodological quality, five studies of medium quality, and three studies of low quality. According to four studies extracorporeal membrane oxygenation did not reduce mortality in adults (OR/RR ranging from 0.71 to 1.28), but two studies reported significantly lower mortality in patients receiving venovenous extracorporeal membrane oxygenation than in the control group (OR/RR ranging from 0.38 to 0.73). Besides, monitoring of vital signs and increasing the number of medical staff may also reduce the mortality in patients with respiratory diseases.

**Conclusions:** Our overview suggests that supportive care may reduce the mortality of patients with respiratory diseases to some extent. However, the quality of evidence for the primary outcomes in the included studies was low to moderate. Further systematic reviews and meta-analyses are needed to address the evidence gap regarding the supportive care for SARS, MERS and COVID-19.

## Background

The aim of supportive care is to prevent or treat the symptoms of a disease, side effects caused by treatment of a disease, and psychological and social problems related to a disease or its treatment as early as possible (1). Supportive care can also be called comfort care, palliative care, or symptom management. Supportive care can improve the quality of life of patients who have a serious or life-threatening disease, such as cancer (2). Besides, for patients with respiratory diseases, supportive care also is an important and effective part of the management (3). A systematic review related to severe acute respiratory syndrome (SARS) treatment suggested that meticulous supportive care is the only form of treatment that can be recommended (4). Another systematic review showed that continuous monitoring of vital signs outside the critical care setting is feasible and may provide a benefit in terms of improved patient outcomes and cost efficiency (5). According to the World Health Organization (WHO) guidelines there is currently no treatment recommended for coronavirus infections except supportive care as needed (6).

In early 2020, a pneumonia caused by a novel coronavirus (SARS-CoV-2) emerged in Wuhan, China, and rapidly spread to more than 100 countries around the world (7). As of 12 April 2020, more than 1,690,000 cases and more than 100,000 deaths have been confirmed according to the WHO (8). The disease caused by SARS-CoV-2, coronavirus disease 2019 (COVID-19), has had been declared a global pandemic (9) However, there is so far no effective treatment or vaccine to curb the spread of the epidemic. Thus, we conducted this overview to identify the available and effective forms of supportive care for patients with respiratory diseases. We hope this review will help physicians working on COVID-19 to understand more about supportive care and make decisions on treatment selection for COVID-19.

## Methods

### Search strategy

We performed a systematic search of Medline via PubMed, EMBASE, Web of Science, CNKI (China National Knowledge Infrastructure), Wanfang Data and CBM (China Biology Medicine disc) from their inception to 31 March 2020 with the terms (“COVID-19” OR “SARS-CoV-2” OR “2019 novel coronavirus” OR “2019-nCoV” OR “Wuhan coronavirus” OR “novel coronavirus” OR “Wuhan seafood market pneumonia virus” OR “MERS” OR “SARS” OR “Severe Acute Respiratory Syndrome” OR “Middle East Respiratory Syndrome Coronavirus” OR “Influenza”) AND (“Meta-analysis” OR “Systematic Review” OR “rapid review”) (The details of the search strategy can be found in the Supplementary Material 1). Search strategies for other databases are adapted from PubMed. In addition, we searched Google Scholar as well as reference lists of the identified articles, to find additional studies. Three preprint services, including medRxiv (https://www.medrxiv.org/), bioRxiv (https://www.biorxiv.org/) and SSRN (https://www.ssrn.com/index.cfm/en/) were also searched to find relevant studies.

### Inclusion and exclusion criteria

We included systematic reviews and meta-analyses related to supportive treatment for patient with respiratory diseases including COVID-19, SARS, MERS and influenza published in English and Chinese without other restrictions. We included systematic reviews and meta-analyses that focused on the proportion of medical staff, monitoring of vital signs, respiratory and circulatory support, and psychological intervention. We also considered systematic reviews and meta-analyses with related indirect evidence if no sufficient literature on COVID-19, SARS, MERS and influenza was found. We excluded duplicates, conference abstracts and articles where we failed to access full text and data despite contacting the authors.

### Study selection and data extraction

Two reviewers (X Luo and M Lv) screened all titles, abstracts and full texts independently and solved disagreements by consensus or consultation with a third reviewer. We extracted the following basic information: 1) title, 2) first author, 3) publication year, 4) journal, 5) number of included studies, 6) study design of included studies, and 7) sample size; and the following information on the results; 1) primary outcome, 2) effect size (odds ratio, OR; relative risk, RR), 3) 95% confidence interval (CI), 4) heterogeneity, and 5) main conclusion.

### Quality assessment

Two researchers (X Wang and X Zhang) independently evaluated the quality of the included studies and cross-checked the results. If necessary, a third reviewer (X Luo) participated in the discussion. Methodological quality assessment of included literature was performed using the AMSTAR tool (10). The AMSTAR score has a total of 11 points, with studies scoring between 9 and 11 being considered to be of high quality, studies scoring between 6 and 8 of medium quality, and studies scoring between 0 and 5 of low quality. We evaluated the quality of evidence for the primary outcomes of each included systematic review and meta-analysis using the GRADE method (11).

### Data synthesis

We conducted a descriptive analysis of the included literature. We analyzed studies on the proportion of medical staff, monitoring of vital signs, respiratory and circulatory support, and psychological intervention for patients with respiratory diseases separately. All statistical analyses were conducted in STATA 14.0. A random-effects model was used to show the primary outcomes from each systematic review and meta-analysis.

## Results

A total of 3536 records were identified. After reading the full texts, eighteen systematic reviews and meta-analyses were included (5,12-28) (*Figure 1*). Twelve reviews (13,14,16-18,22-28) studied respiratory and circulatory support, three (5,15,21) the monitoring of vital signs, two (19,20) the proportion of medical staff, and one (12) psychological impact. (*Table 1*)

**Table 1.**
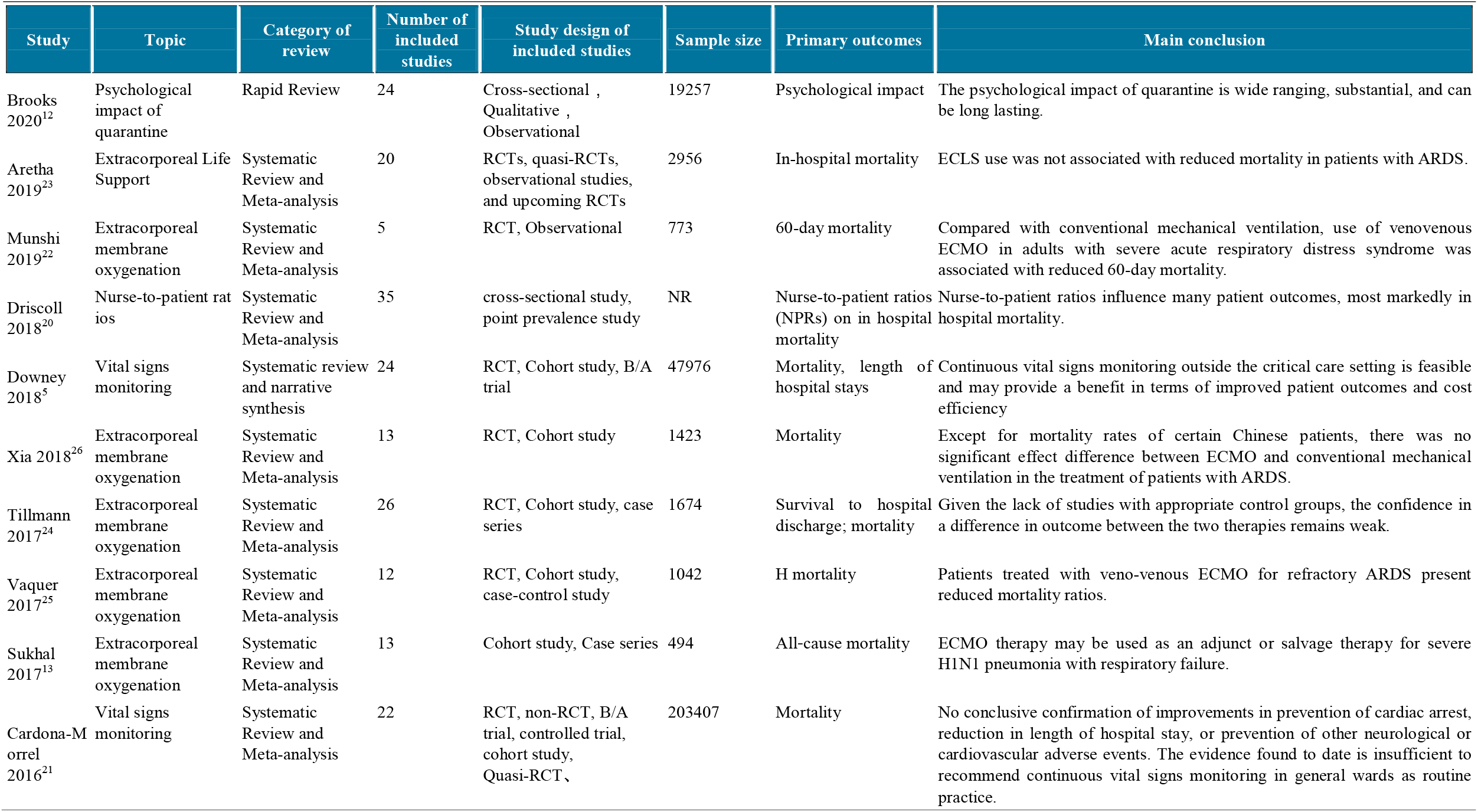
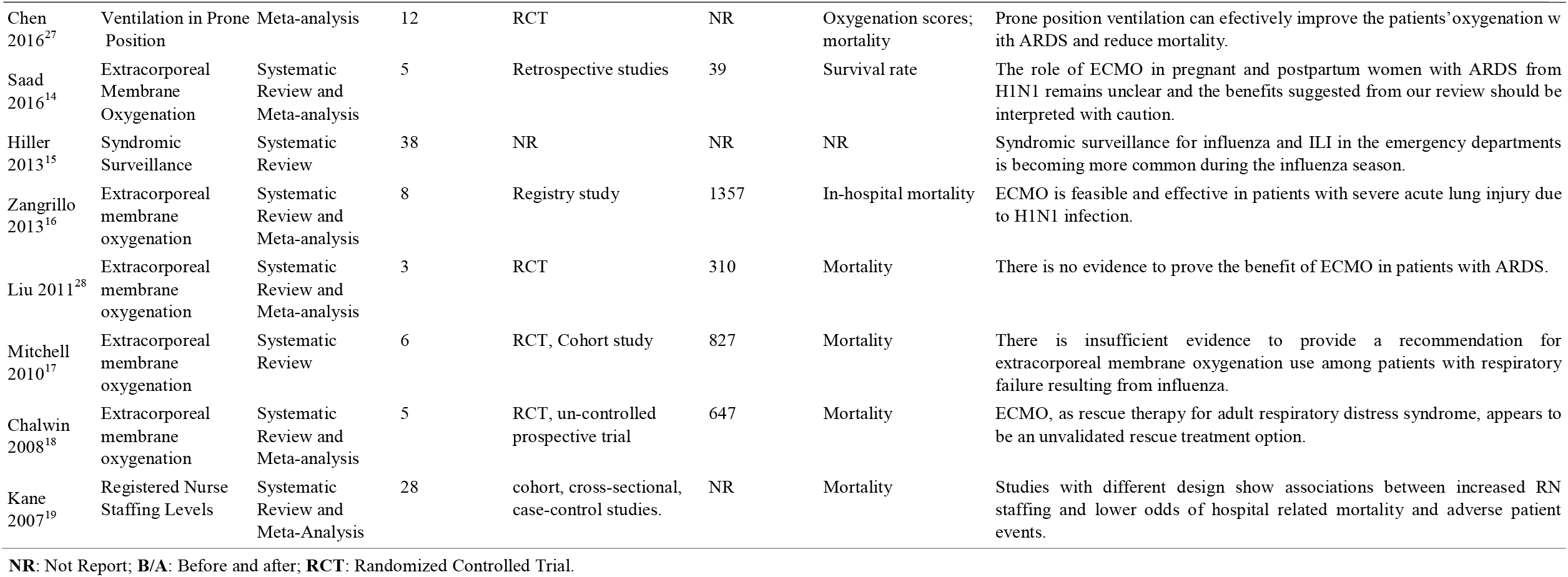
Baseline characteristics of the included studies.

**Figure 1.**
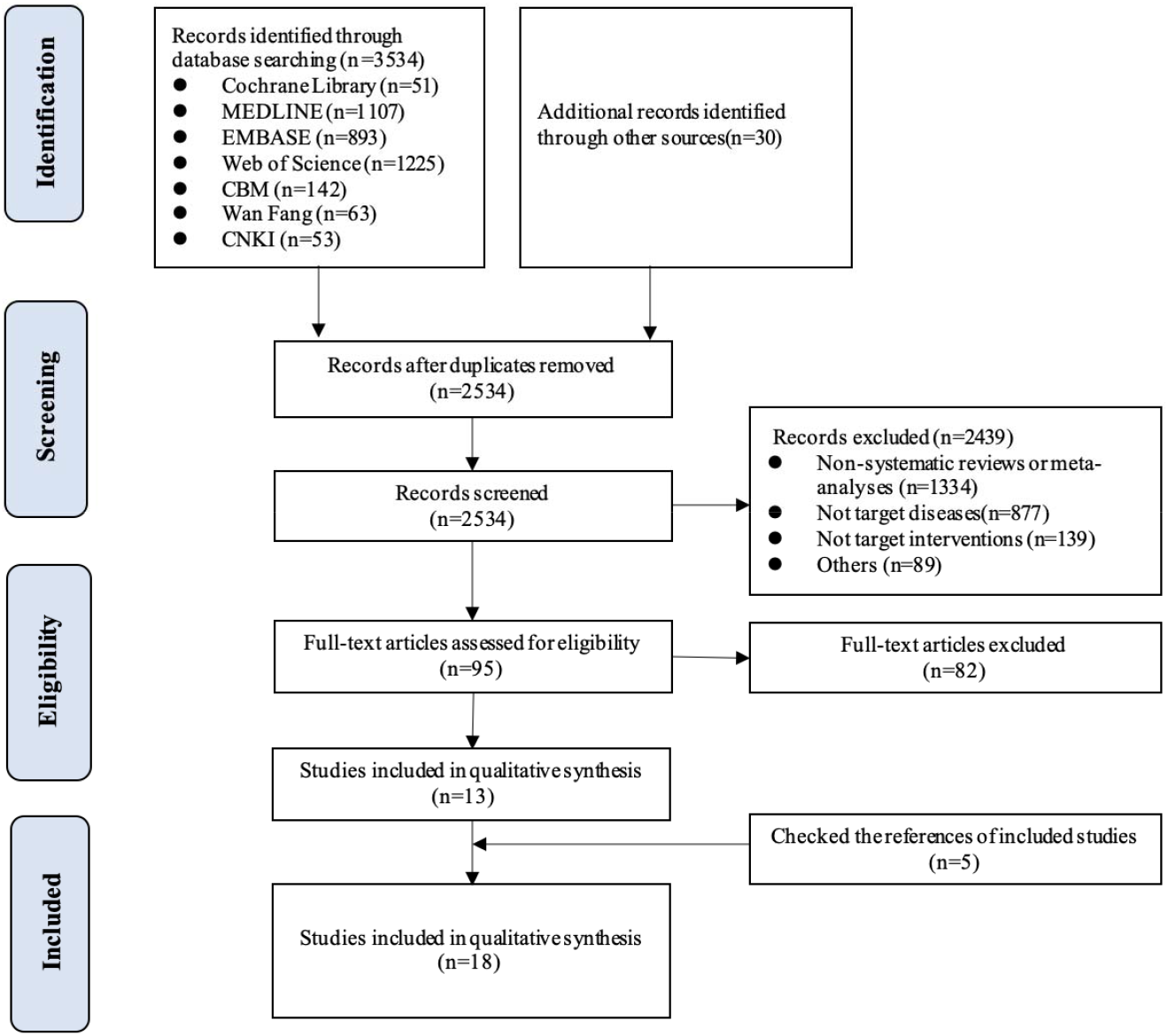
Flow chart of literature screening and selection.

### Quality assessment of included studies

According to the AMSTAR scores, ten studies (13-15,20-26) were of high quality, five studies (5,16,17,19,28) of medium quality, and three studies (12,18,27) of low quality (*Table 2*). According to our assessment using the GRADE approach, five of the 15 primary outcomes were based on moderate-quality evidence, and ten on low-quality evidence (*Table 3*).

**Table 2.**
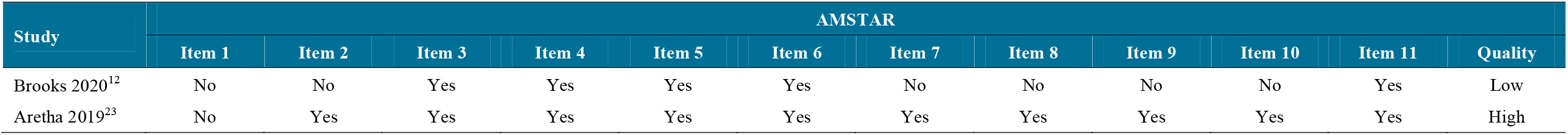
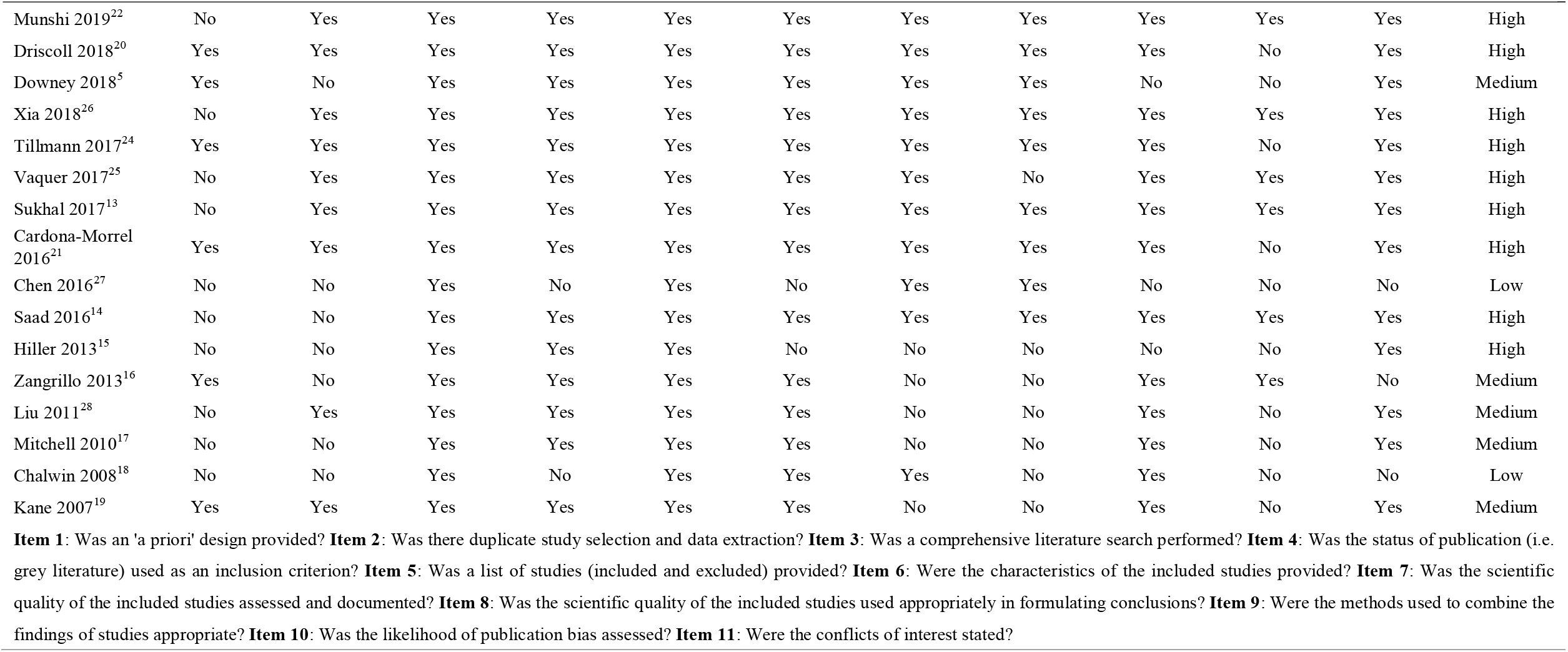
Methodological quality of included systematic reviews and meta-analyses.

**Table 3.** Quality of evidence of primary outcomes in included studies.

| Quality assessment |  |  |  |  |  |  |  | Effect size<br>95% CI | Quality of the<br>evidence<br>(GRADE) |
| --- | --- | --- | --- | --- | --- | --- | --- | --- | --- |
| No. of studies<br>(sample size) | Study design | Risk of bias | Inconsistency | Indirectness | Imprecision | Publication<br>bias | Rating up<br>factor |  |  |
| Extracorporeal Membrane Oxygenation: survival rate |  |  |  |  |  |  |  |  |  |
| 5 (39) | retrospective study | Serious<sup>a</sup> | Not serious | Not serious | Not serious | Undetected | No | ES 0.75<br>[0.61, 0.89] | ⊕⊕⊕○<br>Moderate |
| Extracorporeal Membrane Oxygenation: mortality |  |  |  |  |  |  |  |  |  |
| 8 (1357) | registry study | Serious<sup>a</sup> | Serious<sup>b</sup> | Not serious | Not serious | Undetected | No | ES 0.28 | ⊕⊕○○ |
|  |  |  |  |  |  |  |  | [0.18-0.37] | Low |
| Extracorporeal Membrane Oxygenation: mortality |  |  |  |  |  |  |  |  |  |
| 3 (310) | RCT | Serious<sup>a</sup> | Not serious | Serious<sup>c</sup> | Not serious | Undetected | No | RR 0.95<br>[0.76-1.18] | ⊕⊕OO |
|  |  |  |  |  |  |  |  |  | Low |
| Extracorporeal Membrane Oxygenation: mortality |  |  |  |  |  |  |  |  |  |
| 6 (827) | RCT | Not serious | Serious<sup>b</sup> | Not serious | Not serious | Undetected | No | RR 0.93<br>[0.71-1.22] | ⊕⊕⊕O |
|  |  |  |  |  |  |  |  |  | Moderate |
| Extracorporeal Membrane Oxygenation: mortality |  |  |  |  |  |  |  |  |  |
| 5 (647) | RCT | Not serious | Not serious | Serious<sup>c</sup> | Serious<sup>d</sup> | Undetected | No | OR 1.28<br>[0.24-6.55] | ⊕⊕OO |
|  |  |  |  |  |  |  |  |  | Low |
| Extracorporeal Membrane Oxygenation: mortality |  |  |  |  |  |  |  |  |  |
| 5 (773) | RCT | Serious<sup>a</sup> | Not serious | Not serious | Not serious | Undetected | No | RR 0.73<br>[0.58-0.92] | ⊕⊕⊕O |
|  |  |  |  |  |  |  |  |  | Moderate |
| Extracorporeal Membrane Oxygenation: mortality |  |  |  |  |  |  |  |  |  |
| 20 (2956) | mixed | Not serious | Serious<sup>b</sup> | Serious<sup>c</sup> | Not serious | Undetected | No | OR 0.96<br>[0.57-1.77] | ⊕⊕OO |
|  |  |  |  |  |  |  |  |  | Low |
| Extracorporeal Membrane Oxygenation: mortality |  |  |  |  |  |  |  |  |  |
| 26 (1674) | mixed | Not serious | Serious<sup>b</sup> | Serious<sup>c</sup> | Not serious | Undetected | No | RR 0.71<br>[0.33-1.51] | ⊕⊕OO |
|  |  |  |  |  |  |  |  |  | Low |
| Extracorporeal Membrane Oxygenation: mortality |  |  |  |  |  |  |  |  |  |
| 12 (1042) | mixed | Not serious | Serious<sup>b</sup> | Serious<sup>c</sup> | Not serious | Undetected | No | OR 0.38<br>[0.32, 0.44] | ⊕⊕OO |
|  |  |  |  |  |  |  |  |  | Low |
| Extracorporeal Membrane Oxygenation: mortality |  |  |  |  |  |  |  |  |  |
| 13 (1423) | mixed | Not serious | Serious<sup>b</sup> | Serious<sup>c</sup> | Not serious | Undetected | No | OR 1.12<br>[0.69-1.81] | ⊕⊕OO |
|  |  |  |  |  |  |  |  |  | Low |
| Extracorporeal Membrane Oxygenation: mortality |  |  |  |  |  |  |  |  |  |
| 13 (494) | cohort study | Serious<sup>a</sup> | Serious<sup>b</sup> | Not serious | Not serious | Undetected | No | OR 0.37<br>[0.30-0.45] | ⊕⊕OO |
|  |  |  |  |  |  |  |  |  | Low |
| Registered Nurse Staffing Levels: mortality |  |  |  |  |  |  |  |  |  |
| 28 (NR) | mixed | Serious<sup>a</sup> | Not serious | Not serious | Not serious | Undetected | No | OR 0.91<br>[0.86-0.96] | ⊕⊕⊕O |
|  |  |  |  |  |  |  |  |  | Moderate |
| Nurse-to-patient ratios: mortality |  |  |  |  |  |  |  |  |  |
| 35 (NR) | mixed | Serious<sup>a</sup> | Serious<sup>b</sup> | Not serious | Not serious | Undetected | No | OR 0.86<br>[0.79-0.94] | ⊕⊕OO |
|  |  |  |  |  |  |  |  |  | Low |
| Vital signs monitoring: mortality |  |  |  |  |  |  |  |  |  |
| 22 (203407) | RCT | Not serious | Serious<sup>b</sup> | Not serious | Not serious | Undetected | No | OR 0.83 | ⊕⊕⊕O |
|  |  |  |  |  |  |  |  | [0.53-1.29] | Moderate |
| Ventilation in Prone Position: mortality |  |  |  |  |  |  |  |  |  |
| 12 (NR) | RCT | Not serious | Serious<sup>b</sup> | Serious<sup>c</sup> | Not serious | Undetected | No | OR 0.63<br>[0.51-0.78] | ⊕⊕○○ |
|  |  |  |  |  |  |  |  |  | Low |
| **High:** We are very confident that the true effect lies close to that of the estimate of the effect.<br>**Moderate:** We are moderately confident in the effect estimate: The true effect is likely to be close to the estimate of the effect, but there is a possibility that it is substantially different.<br>**Low:** Our confidence in the effect estimate is limited: The true effect may be substantially different from the estimate of the effect.<br>**Very low:** We have very little confidence in the effect estimate: The true effect is likely to be substantially different from the estimate of effect. |  |  |  |  |  |  |  |  |  |

<sup>a</sup> **Quality of included studies poor because of inadequate study design and follow-up time.**
<sup>b</sup> **Serious inconsistency for the scattered 95% CI.**
<sup>c</sup> **Indirect evidence for target population.**
<sup>d</sup> **Wide confidence intervals, serious imprecision.**

### Respiratory and circulatory support

Twelve included systematic reviews and meta-analyses focused on respiratory circulation support, of which seven studies explored the role of extracorporeal membrane oxygenation (ECMO) in reducing in the mortality of patients with acute respiratory distress syndrome (ARDS), and four studies reported outcomes of ECMO on H1N1. One study explored the effects of prone ventilation on oxygenation index and mortality in patients with ARDS (*Figure 2*).

**Figure 2.**
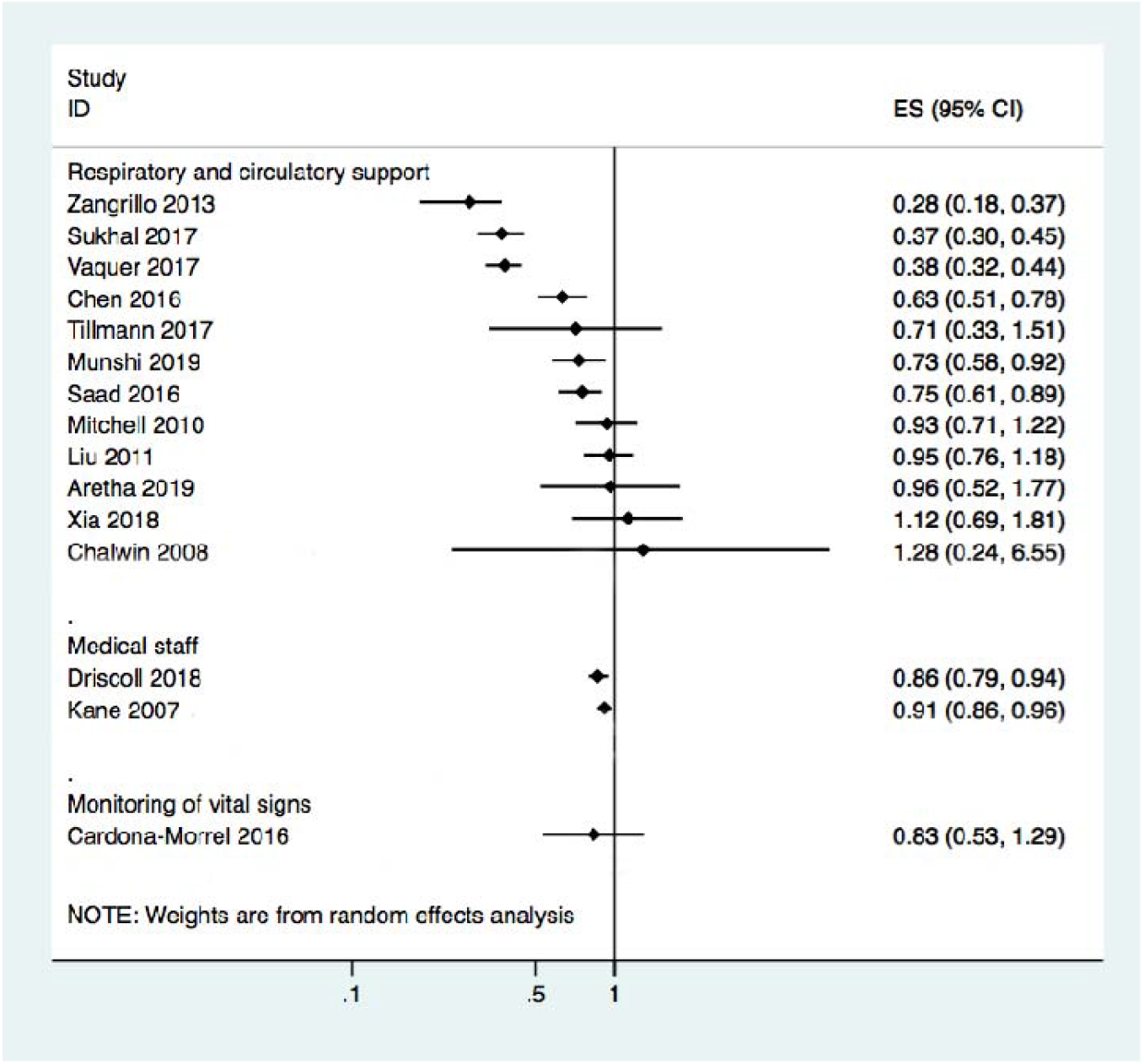
Results of meta-analyses on mortality from the included systematic reviews. The effect sizes are reported either as risk ratio or odds ratio comparing the risk/odds of death in the intervention with control group.

Of the 11 systematic reviews and meta-analyses on ECMO, seven reported the impact of ECMO on ARDS; four of them reviewed the impact of ECMO on ARDS in adults, and the results of the four studies showed that ECMO did not reduce the mortality of adults. There was large heterogeneity between the included studies in the reviews. One study reported that mortality in ECMO group was lower than control group in the subgroup of Chinese patients [OR=0.39, 95% CI, 0.17-0.86]. Another article reported that the probability of experiencing at least one adverse event of ECMO varied greatly between studies, with a range of 1.1%-100%. Two other meta-analyses reported a reduction in ARDS related mortality in patients having venovenous ECMO. Both meta-analyses showed that mortality was significantly lower in the venovenous ECMO group than in the control group. However, both studies also reported an association between venovenous ECMO and bleeding. Finally, one study reported the mortality rate in patients receiving extracorporeal life support (ELS) for the treatment of ARDS. Meta-analysis showed that ELS failed to show any survival benefit in ARDS patients, and the heterogeneity among studies was large; however, when the study type was limited to RCT, there was a mortality difference favoring the ECMO group [OR=0.51, 95% CI, 0.37-0.70, *P*=0.33, *I*^*2*^=12.2%], and bleeding was more common in the ELS group.

Of the 11 studies, four focused on H1N1 and conditions associated with it. Two reviews reported the outcomes of patients infected with H1N1. The results of one meta-analysis showed that outcomes were highly variable among the included studies, with short-term in-hospital risk of death ranging between 8% and 65%, mainly depending on baseline patient characteristics. Random-effect pooled estimates suggested a 28% risk of in-hospital death [95% CI, 18%-37%] among patients with H1N1-associated severe acute lung injury. According to another systematic review, the risk ratio for death with versus without ECMO based on three trials was 0.93 [95% CI, 0.71-1.22],and the heterogeneity among studies was large. One systematic review indicated that the pooled estimate of the survival probability among pregnant and postpartum patients who received ECMO was 74.6% [95% CI, 60.7%-88.6%]. Heterogeneity was not significant in any combination of four of the five included primary studies (*I*^*2*^=0-21%; *P*>0.25). The last of the four reviews reported the outcomes of severe influenza infection with respiratory failure. The overall risk of death was 37% [95% CI, 30%-45%], the median duration for ECMO was 10 days and for mechanical ventilation 19 days, and the median length of ICU stay was 33 days. However, the heterogeneity among studies was large (*I*^*2*^=65%).

One review reviewed the influence of ventilation in prone position on patients with ARDS. Seven of the 12 included studies in this review evaluated the effect of the prone position on the oxygenation index. The oxygenation index of the group receiving ventilation in the prone position was significantly higher than that of the control group (ventilation in supine position) [OR=69.65, 95% CI, 37.13-102.7, *P*<0.001]. Eight studies evaluated the impact of the prone position on the mortality, showing that the mortality in the prone position was significantly lower than that in the supine position [OR= 0.63, 95% CI, 0.51-0.78, *P*<0.001].

### Monitoring of vital signs

Three systematic reviews and meta-analyses explored the impact of monitoring vital signs and symptoms on influenza patients and the general population (*Figure 2*). One systematic review published in 2018 assessed if continuous monitoring is practical outside of the critical care setting, and whether it confers any clinical benefit to patients. The majority of studies showed benefits such as less need for critical care and shorter hospital stay. Larger studies were more likely to demonstrate clinical benefit, particularly in the need of critical care use and length of hospital stay. Barriers to implementation of monitoring of symptoms and vital signs included concerns about the negative effects among nurses and patients, and the burden of false alerts. In summary, continuous monitoring of vital signs outside the critical care setting is feasible and may provide a benefit in terms of improved patient outcomes.

A systematic review and meta-analysis published in 2016 identified strategies to improve the monitoring of intermittent or continuous vital signs and prevent adverse events in general hospital wards. The results suggested that strategies for monitoring continuous vital signs were not associated with significant reductions in in-hospital mortality [OR=0.87, 95% CI, 0.57-1.33]. There was only minor heterogeneity between studies (*I*^*2*^=27%, *P*=0.25). In contrast, enhanced monitoring of intermittent vital signs was associated with modest reduction in the risk of death when compared with usual care [OR=0.78, 95% CI, 0.61-0.99]. However, there was no evidence of reduction in the need of transfer to ICU or in adverse events with either intermittent or continuous monitoring.

Besides, the syndromic surveillance for influenza and influenza-like-illness from the emergency department is becoming more common to detect yearly influenza outbreaks, as shown in a systematic review.

### Medical staff

Two systematic reviews reported the impact of the number of medical staff on disease outcomes (*Figure 2*). One examined the association between registered nurse (RN) staffing and patient outcomes in acute care hospitals. The results showed that increased RN staffing was associated with lower mortality in intensive care units (ICUs) [OR=0.91, 95% CI, 0.86-0.96], in surgical patients [OR=0.84, 95% CI, 0.80-0.89), and in medical patients [OR=0.94, 95% CI, 0.94-0.95]; all ORs reported per additional full time equivalent RN per patient-day). An increase by one RN per patient-day was associated with decreased odds of hospital acquired pneumonia [OR=0.70; 95% CI, 0.56-0.88], unplanned extubation [OR=0.49, 95% CI, 0.36-0.67], respiratory failure [OR=0.40, 95% CI, 0.27-0.59], and cardiac arrest (OR=0.72, 95% CI, 0.62-0.84] in ICUs, with a lower risk of failure to rescue [OR=0.84, 95% CI, 0.79-0.90] in surgical patients. Increase of one RN per patient-day also shortened the length of stay in ICUs [OR=0.76, 95% CI, 0.62-0.94] and in surgical patients [OR=0.69, 95% CI, 0.55-0.86]. Another study examined the association between nurse staffing levels and nurse-sensitive patient outcomes in acute special units. The results of the meta-analysis, comprising of six original studies, reported ORs on all-cause in hospital mortality of 175,755 patients admitted to ICUs or cardiac or cardiothoracic units. A higher level of nurse staffing decreased the risk of in-hospital death by 14% [95% CI, 0.79-0.94]. However, the meta-analysis also showed high heterogeneity (*I*^*2*^=86%).

### Psychological impact

A rapid review published in 2020 focused on the psychological impact of quarantine to explore its likely effects on mental health and psychological wellbeing, and the factors that contribute to, or mitigate, these effects. A total of 24 studies were included and the results showed that stressors during quarantine were 1) duration of quarantine; 2) fears of infection; 3) frustration and boredom; 4) inadequate supplies; and 5) inadequate information. The stressors during post-quarantine time were finances and stigma. This rapid review suggested to keep quarantine time as short as possible, give people as much information as possible, provide adequate supplies, reduce the boredom, and improve the communication. Special attention should be paid on health-care workers’ mental health, and it is advisable to appeal to people’s altruism and the public health benefits of isolation rather than compulsion. Overall, the psychological impact of quarantine is wide-ranging, substantial, and can be long lasting. Officials should quarantine individuals for no longer than required, provide clear rationale for quarantine and information about protocols, and ensure sufficient supplies are provided.

## Discussion

Our study showed that for patients with respiratory diseases, especially H1N1 patients, ECMO may effectively reduce mortality, but attention should be paid to the risk of bleeding. For patients in non-critical wards, monitoring and recording of vital signs can effectively reduce mortality. In addition, increasing the number of medical staff in intensive care units can also reduce mortality. At the same time, psychological intervention is equally important for isolating patients. All systematic reviews were of low to moderate quality.

Supportive treatment is an important and effective part of the management for patients with respiratory diseases (29). After systematic searching, we did not find systematic reviews or meta-analyses for supportive care in SARS, MERS or COVID-19-patients, and only five systematic reviews and meta-analyses for influenza. In the absence of direct evidence, we focused on systematic reviews and meta-analysis of ECMO for ARDS. Overall, compared with the control group, ECMO did not reduce mortality in adult patients with ARDS, and the quality of evidence was relatively low due to the large heterogeneity between studies. ECMO can improve severe hypoxemia in patients with ARDS, keep the lungs at rest, and wait for lung tissue to repair, but the clinical research results on the prognosis of patients with ARDS are not consistent. At the same time, two systematic reviews have shown that venovenous ECMO can reduce ARDS-related mortality, however the risk of bleeding needs to be considered. For patients with H1N1, one of the two currently available systematic reviews also reported that ECMO was associated with reduced risk of death (13,16). A systematic review (16) published in 2013 suggested that ECMO is effective in reducing the mortality of patients with H1N1, but not among patients with ARDS caused by other pathogens. The benefit of ECMO may thus depend on the cause of ARDS (30).

Monitoring vital signs with non-critical patients can reduce patient mortality and shorten the length of hospital stay, but the specific circumstances of the patient need to be considered, such as affordability and acceptability (31).

Increasing the number of medical staff in the intensive care unit can reduce the mortality of patients. The likely reason is that due to the increase in the number of caregivers, patients can receive more time, attention and care. However, given the limited medical resources, investing in more medical staff into the ICU may need to be balanced by savings in some other expenditure, for example staff in other departments. Therefore, investing more medical staff into the ICU still needs further evaluation. In addition, for infectious diseases like SARS, MERS and COVID-19, psychological intervention is equally important. We found systematic reviews that analyzed the factors that may cause mental illness and provides psychological precautions, but none of them considered COVID-19. For psychological interventions on COVID-19, relevant research is still needed.

We used the AMSTAR tool to evaluate the methodological quality of the included systematic reviews and meta-analyses and found that the overall quality is relatively high. In terms of evidence quality, the main problem was the large heterogeneity between included studies. The included studies contained only indirect evidence, and the reliability of the overall quality of evidence was low to moderate, which must be taken into consideration when the evidence is used for making clinical practice guidelines.

To our knowledge, this is the first overview of systematic review focusing on the supportive care for patient with respiratory diseases. We comprehensively searched systematic reviews and meta-analyses on SARS, MERS, COVID-19 and influenza and evaluated the quality of methodology and evidence. Nevertheless, our work has several limitations. First, we conducted a rapid literature searching and screening, and some relevant studies may have been missed. Second, none of the included systematic reviews focused on COVID-19, SARS or MERS, so they can be only used as indirect evidence. Finally, there was large heterogeneity between the included studies, and most primary studies included in these systematic reviews were observational studies, which may influence the reliability of the reviews. However, systematic reviews and meta-analyses of non-randomized studies can be meaningful and guide clinical research and practice, even if only by emphasizing the limitations of the available clinical evidence.

## Conclusion

In conclusion, our overview suggests that supportive cares may reduce the mortality of patients with respiratory diseases to some extent. Having more medical staff in ICU, using of ECMO, monitoring vital signs and conducting counselling to patients exposed to respiratory diseases are all effective measures to improve the patients’ outcomes. However, further studies are needed to address the evidence gap regarding the supportive care for SARS, MERS and COVID-19.

## Data Availability

All relevant data are listed in the full text.

## Author contributions

(I) Conception and design: Y Chen, XH Wang and E Liu; (II) Administrative support: Y Chen; (III) Provision of study materials or patients: X Luo, M Lv and XQ Wang; (IV) Collection and assembly of data: X Luo and M Lv; (V) Data analysis and interpretation: X Luo, XH Wang and M Lv; (VI) Manuscript writing: All authors; (VII) Final approval of manuscript: All authors.

## Acknowledgments

We thank Janne Estill, Institute of Global Health of University of Geneva for providing guidance and comments for our review. We thank all the authors for their wonderful collaboration.

## Footnote

### Ethical Statement

The authors are accountable for all aspects of the work in ensuring that questions related to the accuracy or integrity of any part of the work are appropriately investigated and resolved.

**Supplementary Material 1** Search Strategy for PubMed

#1. “severe acute respiratory syndrome coronavirus 2” [Supplementary Concept]

#2. “COVID-19” [Supplementary Concept]

#3. “Novel coronavirus” [Title/Abstract]

#4. “2019-novel coronavirus” [Title/Abstract]

#5. 2019-nCoV [Title/Abstract]

#6. “Novel CoV” [Title/Abstract]

#7. “Wuhan-Cov” [Title/Abstract]

#8. “2019-CoV” [Title/Abstract]

#9. “Wuhan Coronavirus” [Title/Abstract]

#10. “Wuhan seafood market pneumonia virus” [Title/Abstract]

#11. COVID-19 [Title/Abstract]

#12. SARS-CoV-2 [Title/Abstract]

#13. “novel coronavirus pneumonia” [Title/Abstract]

#14. OR/#1-#13

#15. “systematic review” [Publication Type]

#16. “systematic reviews as topic” [MeSH]

#17. meta-analysis [Title/Abstract]

#18. systematic review [Title/Abstract]

#19. rapid review [Title/Abstract])

#20. “Meta-Analysis” [Publication Type]

#21. “Meta-Analysis as Topic” [Mesh]

#22. “Network Meta-Analysis” [Mesh]

#23. OR/#15-#22

#24. #14 AND #23

